# Synapsin autoantibodies during pregnancy are associated with fetal abnormalities

**DOI:** 10.1101/2022.09.23.22280284

**Authors:** Isabel Bünger, Jakob Kreye, Konstantin Makridis, Markus Höltje, Helle Foverskov Rasmussen, Scott van Hoof, Tim Ullrich, Eva Sedlin, Christian Hoffmann, Dragomir Milovanovic, Friedemann Paul, Jessica Meckies, Stefan Verlohren, Wolfgang Henrich, Rabih Chaoui, Angela Kaindl, Harald Prüss

## Abstract

Anti-neuronal autoantibodies can be transplacentally transferred during pregnancy and may cause detrimental effects on fetal development. It is unclear whether autoantibodies against synapsin-I, one of the most abundant synaptic proteins, are associated with developmental abnormalities in humans. We prospectively recruited a cohort of 263 pregnant women and detected serum synapsin-I IgG autoantibodies in 13.3%. Seropositivity was strongly associated with abnormalities of fetal development including intrauterine growth retardation. This finding indicates that these autoantibodies may be clinically useful developmental biomarkers and/or even directly participate in the disease process, thus being amenable to antibody-targeting interventional strategies in the future.

## Introduction

Synapsins are neuron-specific phosphoproteins essential for neurotransmitter release and synaptic plasticity^1^ by controlling the accessibility of synaptic vesicles for exocytosis via interactions with presynaptic proteins and the actin cytoskeleton.^2^ In mammals, three synapsin (SYN) genes *SYN1, SYN2*, and *SYN3* have been identified.^3^ They encode ten isoforms with differentially regulated expression during neurodevelopment that coordinate neurite outgrowth and synapse formation.^4^ Moreover, *Syn1* or *Syn2*, but not of *Syn3* mutant mice displayed a severe epileptic phenotype with generalized seizures that manifested around two to three months of age.^5^ In line with these models, variants in *SYN1* were linked to an X-linked human phenotype comprising epilepsy, learning difficulties, macrocephaly, and aggressive behavior.^6^ Intriguingly, autoantibodies to synapsin-Ia and -Ib have previously been identified in patients with limbic encephalitis and with various psychiatric and neurological disorders.^7^ Despite the location of synapsin-I at the cytoplasmic site of synaptic vesicles, these patient-derived autoantibodies can reach their target via FcγII/III-mediated endocytosis and promote a reduction of synaptic vesicle density, thereby mimicking the human *SYN1* loss-of-function phenotype.^8^ While autoantibodies to synapsin and many further neuronal targets are increasingly detected in neurological disorders,^9^ there is growing evidence that antineuronal autoantibodies can act not only in the individual of their detection, but also in a fetus or neonate when diaplacentally transferred from their mother. In the gestational phase, the active placental transport of IgG antibodies, together with the yet premature fetal blood-brain barrier, may further augment the autoantibody exposure of the developing brain and thereby possibly result in long-lasting neuropsychiatric morbidity.^10^ Such detrimental effects of diaplacentally transferred antineuronal autoantibodies have previously been shown in experimental transfer models of human autoantibodies against N-methyl-D-aspartate (NMDA) receptors,^11^ contactin-associated protein 2 (CASPR2),^12^ aquaporin 4,^13^ and acetylcholine receptors.^14^ In contrast, developmental abnormalities caused by diaplacentally transferred synapsin-I autoantibodies were not studied so far.

Here, we investigated, in an exploratory study, the prevalence of synapsin-I autoantibodies in pregnant women and analyzed their association with fetal development.

## Methods

### Ethics and data protection

Written informed consent was received from all participants prior to study inclusion. Analyses were approved by Charité ethics committee (#EA2/220/20). Samples were handled with pseudonymized identifiers and investigators blinded to the status of donor and fetus.

### Clinical data

Pregnant women presenting for antenatal care in the involved prenatal diagnostic centers between June 2021 and November 2021 were recruited. Medical information was acquired by questionnaire, interview, and from medical documentation. Controls consisting of non-pregnant healthy donors and clinically isolated syndrome (CIS)/relapsing-remitting multiple sclerosis (RRMS) patients were recruited via the DZNE Berlin and the neuroimmunology trial facility at the NeuroCure Clinical Research Center of Charité (NCT01371071, clinicaltrials.gov), respectively. Statistical analysis was performed using Chi-square and Fisher’s exact test, as appropriate.

### Cell-based assay

Synapsin-Ib cell-based assay (CBA) was performed as previously described.^7^ In brief, human embryonic kidney (HEK293T) cells transiently transfected with human synapsin-Ib were methanol-fixed, incubated overnight at 4°C with sera diluted 1:300 and IgG binding detected with a goat anti-human IgG AF488-antibody (Dianova, #109-545-003). For co-stainings, commercial rabbit synapsin-I/II antibody (Synaptic Systems, #106002) and anti-rabbit IgG AF594-antibody (Jackson IR, #111-585-003) were used. The CBA was scored using a semi-quantitative scale by two independent investigators: 0 indicated no binding, 1 unspecific signals (‘background’), and 2 intensive binding (‘positive’) (Fig. 1A). To exclude non-specific HEK293T cell binding, all sera were tested on control cells overexpressing the NR1 subunit of the NMDA receptor or CASPR2, and synapsin CBA-positive sera were also tested on untransfected HEK293T cells.

**Figure 1.**
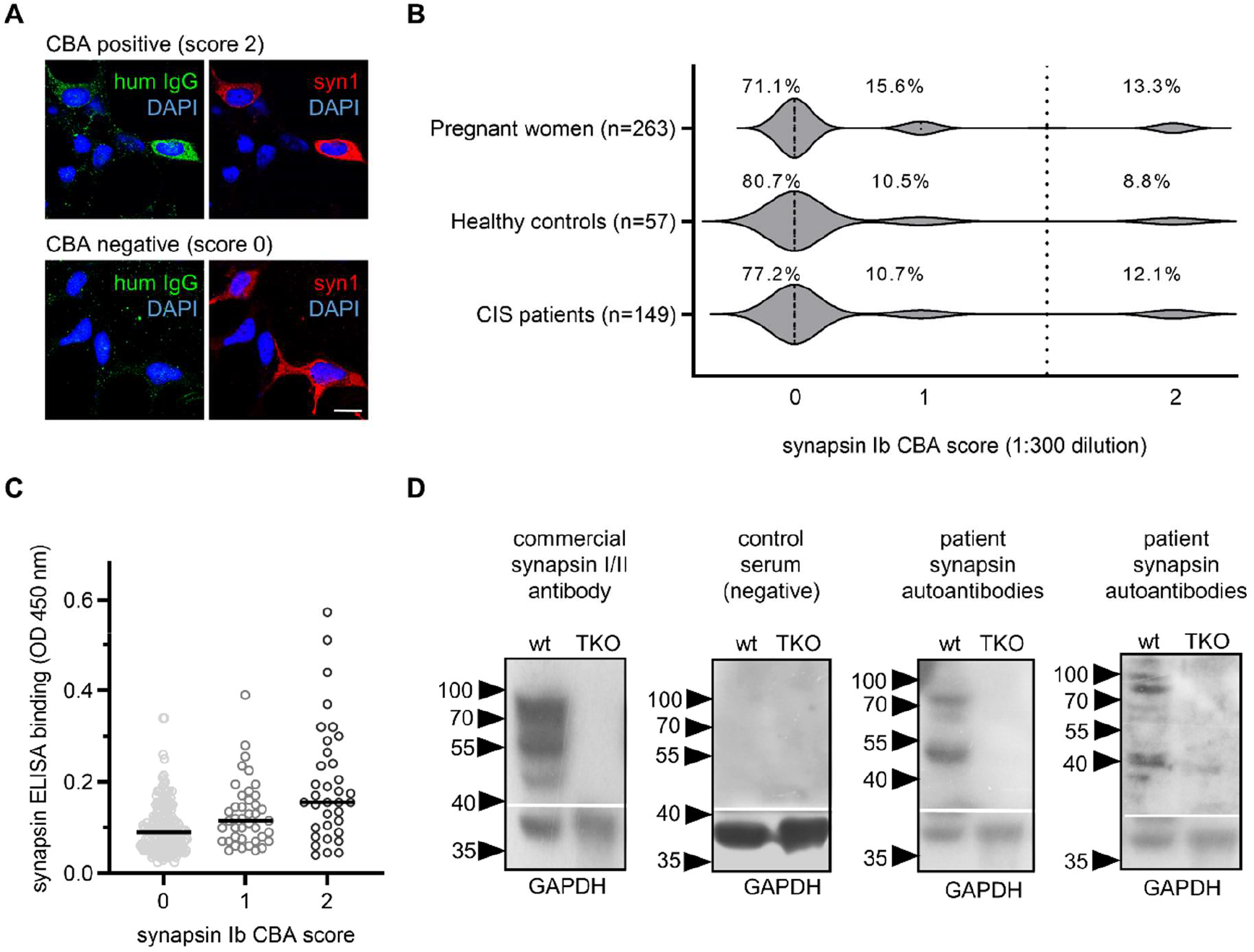
Detection of synapsin I autoantibodies. **(A)** Representative examples of immunofluorescence stainings with human sera at 1:300 dilution (green) with (top row) or without binding (bottom row) to HEK cells overexpressing human synapsin Ib. Protein expression is confirmed with a commercial synapsin I/II antibody (red). Nuclei are stained with DAPI (blue). Scale bar = 20 µm. **(B)** Frequencies of synapsin Ib autoantibodies in pregnant women and control cohorts, as determined by CBA. CBA scores: 0 = no binding, 1 = unspecific signals, 2 = intensive binding (positive); vertical dotted line represents cut-off for positivity. **(C)** Despite higher mean serum IgG levels to immobilized synapsin I (in-house ELISA) in the CBA-positive group, both antibody detection methods did not correlate. **(D)** Representative immunoblots of wild type (wt) and *SynI/II/III* TKO mice cortex homogenates with a commercial synapsin I/II antibody as positive control, and with sera (1:200 dilution) from a CBA-negative control and two CBA-positive pregnant women. Major bands at ∼90 kDa corresponding to the molecular weight of synapsin Ia/Ib and ∼50 kDa that could represent the synapsin IIb isoform or breakdown products of synapsin I were detected in wild type but not in TKO mouse tissue. Detection of GAPDH (Merck Millipore, #MAB374) served as loading control.

### Enzyme-linked immunosorbent assay (ELISA)

Recombinant rat EGFP-synapsin-I^15^ (200 ng) diluted in PBS were incubated overnight at 4°C in 96-well high-binding plates. After blocking, sera diluted 1:200 in blocking solution (PBS containing 1% BSA, 0.05% Tween) were incubated for one hour before adding an HRP-coupled anti-human IgG antibody (Dianova, #109-035-003) for one hour and developed using Ultra TMB substrate solution. 450 nm absorbance values were corrected by subtraction of 630 nm absorbance and by a mean value of control wells containing no serum.

### Western blotting

Cortices from wild type or *Syn1/2/3* triple knockout mice (TKO) were homogenized in RIPA buffer, lysates used for SDS-PAGE and Western blotting as described.^7^ Membranes were incubated with sera of CBA scores 1-2 diluted 1:200. A rabbit polyclonal synapsin-I/II antibody (Synaptic Systems, #106002) served as positive control.

## Results

Given the crucial role of synapsin-I in neurodevelopment, we aimed to screen for synapsin-I autoantibodies during pregnancy and to investigate their association with fetal development. We collected sera from 263 pregnant women (34 ± 5 years of age, 25 ± 8 weeks of gestation; mean ± SD) as well as clinical data (Table 1). Following an explorative approach, there were no exclusion criteria for the recruitment of pregnant women.

**Table 1.** Clinical Data.

|  | Synapsin Ib<br>CBA positive<br>(n=35) | Synapsin Ib<br>CBA negative<br>(n=228) | P value* |
| --- | --- | --- | --- |
| Mother |  |  |  |
| Age (years) | 34 (±5) | 33 (±4) | |
| Chronic diseases | 10 (28.6%) | 42 (18.4%) | 0.160 |
| Autoimmune diseases<sup>a</sup> | 3 (8.6%) | 23 (10.1%) | 1.000 |
| Neuropsychiatric diseases<sup>b</sup> | 3 (8.6%) | 10 (4.4%) | 0.391 |
| Current medication | 11 (12.6%) | 76 (33.3%) | 0.824 |
| Immunomodulatory drugs<sup>c</sup> | 2 (5.7%) | 6 (2.6%) | 0.289 |
| History of current pregnancy |  |  |  |
| Gestational week | 25 (±8) | 25 (±8) | |
| Ultrasound abnormalities | 12 (34.3%) | 33 (14.5%) | **0.004** |
| Structural abnormalities<sup>d</sup> | 8 (22.9%) | 9 (3.9%) | **<0.001** |
| CNS abnormalities<sup>e</sup> | 1 (2.9%) | 6 (2.6%) | 1.000 |
| Growth retardation<sup>f</sup> | 6 (17.1%) | 7 (3.1%) | **0.003** |
| Macrosomia | 1 (2.9%) | 5 (2.2%) | 0.579 |
| Amniotic fluid disorders<sup>g</sup> | 4 (11.4%) | 7 (3.1%) | **0.044** |
| Infections during pregnancy<sup>h</sup> | 2 (5.7%) | 5 (2.2%) | 0.235 |
| Pregnancy-related diseases<sup>i</sup> | 7 (20.0%) | 51 (22.4%) | 0.753 |
| Previous pregnancies |  |  |  |
| Gravida | 2 | 2 |  |
| Para | 1 | 1 |  |
| Miscarriages | 0 | 0 |  |
| Complications during previous pregnancy or labor | 7 (20.0%) | 32 (14.0%) | 0.355 |
| Children with neuropsychiatric disorders | 7 (20.0%) | 15 (6.6%) | **0.016** |

First, all sera were screened for synapsin-Ib autoantibodies of IgG isotype using a previously established CBA.^7^ In the visual scoring system for antibody binding, scores of 2 were considered CBA-positive (Fig. 1A) and detected in 35 (13.3%) pregnant women of our cohort. Similar prevalences were observed in control cohorts of healthy non-pregnant donors (8.8%) and CIS patients (12.1%) (Fig. 1B). Using an alternative detection approach, we analyzed all sera of the pregnant women in an in-house synapsin-I ELISA. Although the group of CBA-positive sera showed more intense binding to the immobilized recombinant synapsin-I, no correlation was detectable between antibody levels in CBA and ELISA (Fig. 1C). One possible explanation for the difference can be antibody binding to conformational epitopes that are present in the CBA but not the ELISA. To this end, we performed immunoblots using the sera and cortex homogenates of wild type and *SynI/II/III* triple TKO mice. Of 35 CBA-positive sera, eight showed also IgG binding to wild type mouse brain homogenates (Fig. 1D), while 27 showed no immunoreactive bands at the expected molecular weight. Thus, IgG autoantibodies likely bind to linear synapsin epitopes only in a minority of those individuals, while autoantibodies to conformational synapsin epitopes may prevail.

Next, we analyzed the clinical data from the women’s pregnancy courses and the fetal development. While there were no differences in the women’s age, gestational age, preexisting conditions, and medication between the two groups, we identified several abnormalities to be significantly associated with the presence of synapsin-Ib autoantibodies (Table 1, Fig. 2). In CBA-positive women fetal ultrasound revealed abnormal results more commonly than in CBA-negative women (p=0.004). Likewise, fetal structural changes were more common in the CBA-positive group (p<0.001). Although developmental changes in the central nervous system were similarly common in both groups, the fetuses of the CBA-positive group displayed an increased frequency of intrauterine growth retardation (p=0.003) and amniotic fluid disorders (p=0.044). Moreover, we found that CBA-positive women more frequently had further children with established neuropsychiatric disorders (p=0.016).

**Figure 2.**
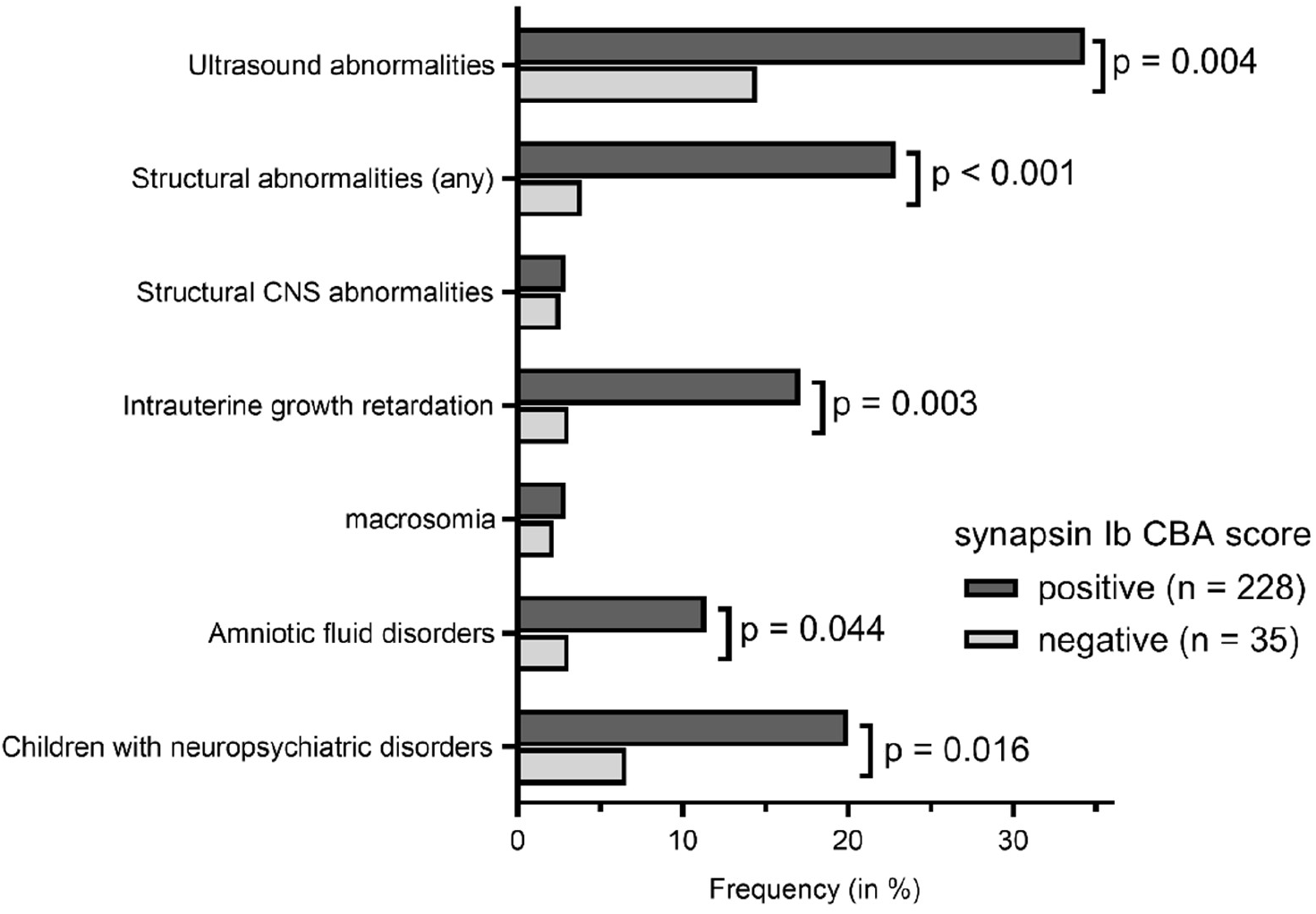
Abnormalities of fetal development associated with synapsin I autoantibodies. Several ultrasound parameters and status of having previous children with neuropsychiatric disorders were significantly associated with the presence of synapsin I autoantibodies in serum of pregnant mothers. Statistical analysis used Fisher’s exact test when expected frequencies were <5 or Chi-square test with expected frequencies ≥5.

## Discussion

In this study, we prospectively recruited a clinical cohort of pregnant women and provide first findings on synapsin-I autoantibodies during pregnancy as well as associations with fetal development. Using a CBA, synapsin-I IgG autoantibodies had a seroprevalence of 13.3% during pregnancy. Most of these cases were negative on Western blots under denaturing conditions, suggesting a conformational dependency of their target binding, a phenomenon known from other human autoantibodies including those against NMDA receptors.^16^ Even though the frequency of high-level synapsin autoantibodies in pregnancy was in the same range as in our two control cohorts, it clearly surpasses frequencies of IgG autoantibodies to other neuronal targets as reported in healthy donors (<2% for 9 tested antigens)^17^ and in mothers of children with neuropsychiatric deficits (4.1% for CASPR2).^18^ Blinded clinical analyses in the present study showed that synapsin-I autoantibodies were strongly associated with several abnormalities of fetal development including intrauterine growth retardation, indicating that the autoantibodies can be a clinically useful biomarker of abnormal fetal development or even directly participate in the disease process. Extensive clinical follow-ups will disclose whether children from seropositive mothers will develop a specific phenotype. Additionally, further studies should investigate synapsin autoantibodies in other clinical cohorts, in particular in children with established neurodevelopmental disorders and their mothers.

While previous experimental data could show that synapsin autoantibodies can reach their intracellular target through endocytosis and cause pathogenic effects *in vitro*,^8^ their role on neurophysiological function and neurodevelopment after transplacental exposure is unclear. Experimental materno-fetal transfer models using antineuronal autoantibodies to other targets^11-13^ have recently substantiated a broader disease principle of neuropsychiatric morbidity resulting from autoantibody exposure during pregnancy. Similar studies are needed for synapsin autoantibodies and should preferentially be performed with patient-derived monoclonal antibodies^19^ to exclude possible interference with other serum antibodies. These experiments will help to elucidate whether synapsin autoantibodies can be directly pathogenic *in vivo*, which seems plausible given that synapsins constitute ∼9% of the total synaptic vesicle proteins^20^ and play a key role in brain development and synaptogenesis. Although the experimental work together with refined epidemiological studies needs to be completed, the present data already support speculation on how transplacental synapsin autoantibodies could change clinical algorithms. This may not only include routine autoantibody screening during or even before pregnancy, but also the development of antibody-targeting interventional strategies, potentially reducing lifelong neuropsychiatric morbidity.

## Data Availability

All data produced in the present study are available upon reasonable request to the authors

## Abbreviations

CASPR2: contactin-associated protein-like 2
CBA: cell-based assay
ELISA: Enzymelinked immunosorbent assay
HEK: Human embryonic kidney
NMDA: N-methyl-Daspartate
SYN: synapsin
TKO: triple knockout mice

## Acknowledgements

We thank Stefanie Bandura, Matthias Sillmann, Doreen Brandl, Antje Dräger, and Monika Majer for excellent technical assistance and study nurse support. J.K. is participant in the Berlin Institute of Health (BIH)-Charité Junior Clinician Scientist Program. This work was supported by grants from the German Research Foundation (DFG) (grants FOR3004, PR1274/4-1, PR1274/5-1, PR1274/9-1), by the Helmholtz Association (HIL-A03), and by the German Federal Ministry of Education and Research (Connect-Generate 01GM1908D) to H.P. The study was supported by the Einstein Stiftung Fellowship through the Günter Endres Fond and the Sonnenfeld-Stiftung (A.M.K.). DM is supported by the start-up funds from DZNE and the German Research Foundation (SFB MI 2104 and 1286/B10). Synapsin triple KO mice were a generous gift from Fabio Benfenati (Istituto Italiano di Tecnologia, Genova).

## Author Contributions

Conceptualization: A.K. and H.P.; Patient recruitment (pregnant women and healthy controls): I.B., K.M., T.U., E.S., J.M., S.V., W.H. and R.C.; Patient recruitment (CIS patients): F.P.; CBA and ELISA testing: I.B., J.K., H.F.R., S.v.H., M.H., C.H. and D.M.; Western blotting M.H.; Data analysis and visualization: I.B. and J.K.; Resources: A.K. and H.P.; Writing – original draft: I.B., J.K. and H.P.; Writing – review & editing: all authors.

## Conflicts of Interest

Nothing to report.

## Notes

### Competing Interest Statement

The authors have declared no competing interest.

### Author Declarations

Ethics Committee of the Charite Universitaetsmedizin Berlin gave ethical approval for this work (#EA2/220/20).

